# Dengue Fever Distribution, Climate and Visitors: A Study from Badung District of Bali, Indonesia

**DOI:** 10.1101/2021.11.15.21266345

**Authors:** Mochamad Rizal Maulana, Ririh Yudhastuti, Muhammad Farid Dimjati Lusno, Yudied Agung Mirasa, Setya Haksama, Zida Husnina

## Abstract

As a national and international tourism destination, Badung district has recorded the highest dengue fever (DF) in Bali Province with multiple suspected influencing factors. This research presents the distribution of dengue fever (DF) incidence in Badung district and analyses its association with climate and visitors. The monthly data of DF surveillance, climate and reported number of visitors during January 2013 – December 2017 was analysed using Poisson Regression. 10689 new DF cases were notified from January 2013 to December 2017 while 2016 was recorded as the heaviest DF incidence. Monthly DF cases have positive association with average temperature, precipitation, humidity and local visitors. While negative association was shown between DF cases with foreign visitors. This study underlines the urgency to integrate climate and tourism for dengue fever control. Further study is recommended to take both factors together with population mobility at finer spatial and time unit of analysis.

## 1. Introduction

The incidence of Dengue Fever (DF) around the world increased in recent decades (Waggoner et al. 2016). The world health organization (WHO) recorded an increasing number of DF cases 15 fold over the last two decades, from 505,430 cases in 2000 to 2,400,138 cases in 2010 and 3,312,040 in 2015. The mortality number increased from 960 in 2000 to more than 4032 in 2015 (WHO 2019). It is estimated 390 million dengue infections occur annually with 96 million cases were manifesting clinical severity (Bhatt et al. 2013). The infection impacted 3.9 billion population in 129 countries at risk, whereas 70 percent of the actual burdens is located in Asia (Brady et al. 2012). Where this region was detected to have profound synchronous dengue fever infection pattern since 1990’s (Van Panhuis et al. 2015).

Dengue fever is caused by *Flaviviridae* family viruses that are categorized into four types (DENV-1, DENV-2, DENV-3, and DENV-4), that all of these strains are exist in Indonesia. The World Health Organization (WHO) classifies Indonesia as one of the endemic countries due to the high periodic DF outbreaks. The DF IR in Indonesia was fluctuated during 2010 – 2017. The lowest IR recorded was 27.67 per 100,000 population (in 2011) and reached the peak 78.85 per 100,000 population in 2017. In 2018, DF incidence was reported by 85.65 percent of all districts in Indonesia. Regions with the highest DF contracted districts were Papua, North Maluku, Bali, Banten, Central Java, Yogyakarta, Jakarta, Jambi, and Riau. Up to 2018, the national IR was IR 24.73/100,000 population with Case Fatality Rates (CFR) was 0.71. This numbers are lower than 2017 with IR 26.10/ 100,000 population with CFR 0.72 percent (MOH 2018).

Bali Province is an endemic are to DF for years. In 2016, the disease transmission emerged in all districts with 21,668 new cases (IR 544.3/100,000 population) and 63 mortalities (CFR 0.29 percent). Badung is the district with the highest number of cases (3,998 cases), followed by Buleleng and Gianyar (3,787 and 3,673 new cases respectively) (PHO 2016). This district has various tourists destinations and has the largest visitors in the Island of Bali. In 2013 as many as 438 thousand local tourists and 3.3 million foreign tourists were recorded to have had visited Badung, and it is expected to increase annually (The Government 2018). This condition was indicated could increase the risk of DF transmission abroad (Yoshikawa and Kusriastuti 2013).

Climate affects the pattern of infectious diseases because vector and viruses sensitivity for any change that directly or indirectly affect the onset of a disease. Some studies have revealed that there is a significant association between DHF with climatic conditions including air temperature, humidity, and precipitation due to increasing breeding place for of *Aedes aegypti* as primary vector of dengue virus (Colón-González et al. 2013, Fatmawati and Sulistyawati 2019, Xiang et al. 2017). Thus, investigation on how the climate variables impact to dengue incidence in Badung could enrich the discussion around this issue.

One of the social demographics and population factors is tourism that have long been studied as the contributing factor to the transmission of infectious disease, as reported by study in South Korea that international travellers from South East Asia are associated with the DF occurence during 2006 – 2015 (Cho et al. 2018). Another dengue case found from traveller who came from Saudi Arabia whose type-2 virus strain clustered with isolates of Singapore and India (Matsui et al. 2019). The impact of mobility and tourism not only exacerbated local transmission but also have driven global spread of DF, for instance the research in 2018 found significant association between the DF incidence in Bali with the imported cases in Australia (Xu et al. 2018).

Despite the outgrowing research around dengue transmission and the high burden of the disease in Badung district, little is known about the distribution characteristics and the association with driving forces both from climate and visitors. Most of the current research examined the association of dengue events with climate factors at one certain time. Such as the study in Denpasar City examined the association of climate and DHF in 2010-2015 (Wiradarma and Somia 2015) another study did comprehensive analysis between DF cases and climate for the entire of Bali Island, however has not taking into account for other factors such as socio economic, transportation and population dynamics (Dhewantara et al. 2019).

Aside from the substantial information presented by these studies, further analysis of DF in relation with climate and visitors at longer observation period could comprehend the recommendation for mitigation and control of DF. In this research, the authors did exploration for the pattern of DF incidence and the explanatory variables (climate and visitors) in Badung district and identify the association between them.

## 2. Materials and Methods

### 2.1 Study design

This research is ecological study with population level as unit analysis. Yearly data from 2013 to 2019 (7 years) at sub-district level was used for descriptive choropleth maps and monthly data for the period of January 2013 – December 2017 (60 months) at district level was used for statistical analysis.

### 2.2 Study Area

Badung district is located between 08°14’20”-08°50’48” South latitude, and 115°05’00”-115°26’16” East longitude, sized 418.5 Kilometers square of lowland area with elevation up to 2075 meters above sea level. Administratively, the district is consisted of 6 sub districts (Petang, Abiansemal, Mengwi, Kuta Utara, Kuta and Kuta Selatan). As tropical area, Badung District has only rainy season and dry season. The rainfall is ranging between 2.0 mm – 622.8 mm. While, the maximum temperature is 26.6 °C – 31.2 °C, the minimum temperature is 25.4 °C - 24.1 °C. The humidity is ranging from 78 percent (around August) – 84 percent (around June).

### 2.3 Variables and Data Sources

The outcome variable in this study is number of DF cases obtained from monthly province surveillance data since January 2013 until December 2017. Dengue fever (DF) is defined as diagnosed by physician with symptoms of 2 – 7 days fever accompanied by manifestations of bleeding, decreased platelets, and hemoconcentration. The confirmation of DF case is supported by Tourniquet test, thrombocytopenia (platelets ≤ 100000/mm3), increased hematocrit ≥ 20 percent, and the increase of the specific IgM antibodies dengue virus in blood tests in the laboratoris (WHO 2011). The independent variables are climate factors (temperature, precipitation and humidity) gathered from Meteorology, Climatology and Geophysics Bureau (BMKG) and number of recorded visitors (local and foreign) were extracted from Badung Statistics Bureau (BPS). The data for producing descriptive mapping was extracted from Badung District health profile 2013 – 2019 that provided DF yearly data at sub-district level categorized based on the gender of reported patients.

### 2.4 Data Analysis

#### 2.4.1 Spatial Visualisation

Choropleth maps were generated to provide geographical distribution of DF incidence rates (IR) across 6 sub-districts for 7 years (2013 – 2019). The incidence rates (IR) was calculated by dividing total cases with the total population of each district and each observed year. The value was classified using natural breaks due to small spatial units. The maps were overlaid with the presentation of gender distribution of reported cases for each sub-districts. The production of maps used ArcGIS 10.4 software by ESRI.

##### Statistical Descriptive and Time Series Decomposition

The descriptive analysis was performed by presenting the variables in distribution frequency table and box plots in order to describe the fluctuation of mean values of each variable in each consecutive years. The DF incidence rates (IR) data was decomposed using multiplicative approach to examine the trend, seasonal and random pattern. The decomposition graph was generated using R statistical software version 4.0.2.

#### 2.4.2 Analysis with Poisson Regression

Uni-variate analysis of Poisson regression was performed to examine the association of DF cases as outcome variable with each of explanatory variables (air temperature, temperature, and precipitation, local and foreign visitors). The analysis was performed for multiple lags variables (1-3 months) to explore the most significant association and to choose variables to be included into the next analysis. The detection for Variance Inflation Factor (VIF) was conducted to examine any multicollinearity the analysis was conducted using STATA 14 software by Stata Corp.

The Multivariate Poisson regression was adopted to examine the association of monthly cases with the selected explanatory variables simultaneously.

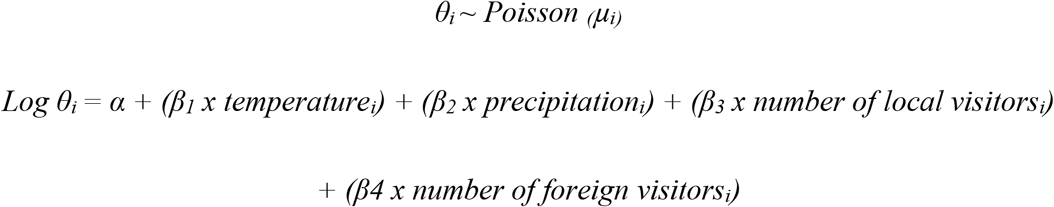

*θ*_*i* j_ is the number of DF cases in Badung district in month i, while *α* is the intercept, *β*_*1*,_ *β*_*2*,_ *β*_*3*_ and *β*_*4*_ are the poisson regression coefficients for the four explanatory variables (temperature, precipitation, number of local visitors and number of foresign visitors). The goodness and significance of the association was evaluated by looking at p value (<0.05), 95% confidence interval (CI) and Akaike Information Criterion (AIC).

##### Ethics

This study was approved by Ethical Committee for Health Research, Faculty of Public Health, Universitas Airlangga, Indonesia, registered on October 1^st^, 2019 with reference number: 208/EA/KEPK/2019.

## 3. Results

### 3.1 Geographics Distribution

In 2016 was recorded as the heaviest DF incidence rates in Badung District where the highest IRs were located in Abiensemal and Mengwi (almost reached 1000 cases/ 100000 population) followed by Kuta Selatan Sub-district. The year of 2015 was recorded as the second highest IR (reached above 500 cases/100000 population) where 2 sub-districts (Mengwi and Kuta Selatan) were the largest transmission. Even though during 2017 and 2018 the IRs were tend to decrease for all 6 sub-districts in Badung District and landed at the lowest in 2018 (below 100 cases/ 100000 population for entire sub-districts). It is however showed darkening IR for Mengwi sub-districts in 2019. Overall, since 2013 – 2019, the areas that frequently experiencing high IR were Mengwi and Kuta Selatan, while the lowest IR were Petang and Kuta district [Figure 1].

**Figure 1:**
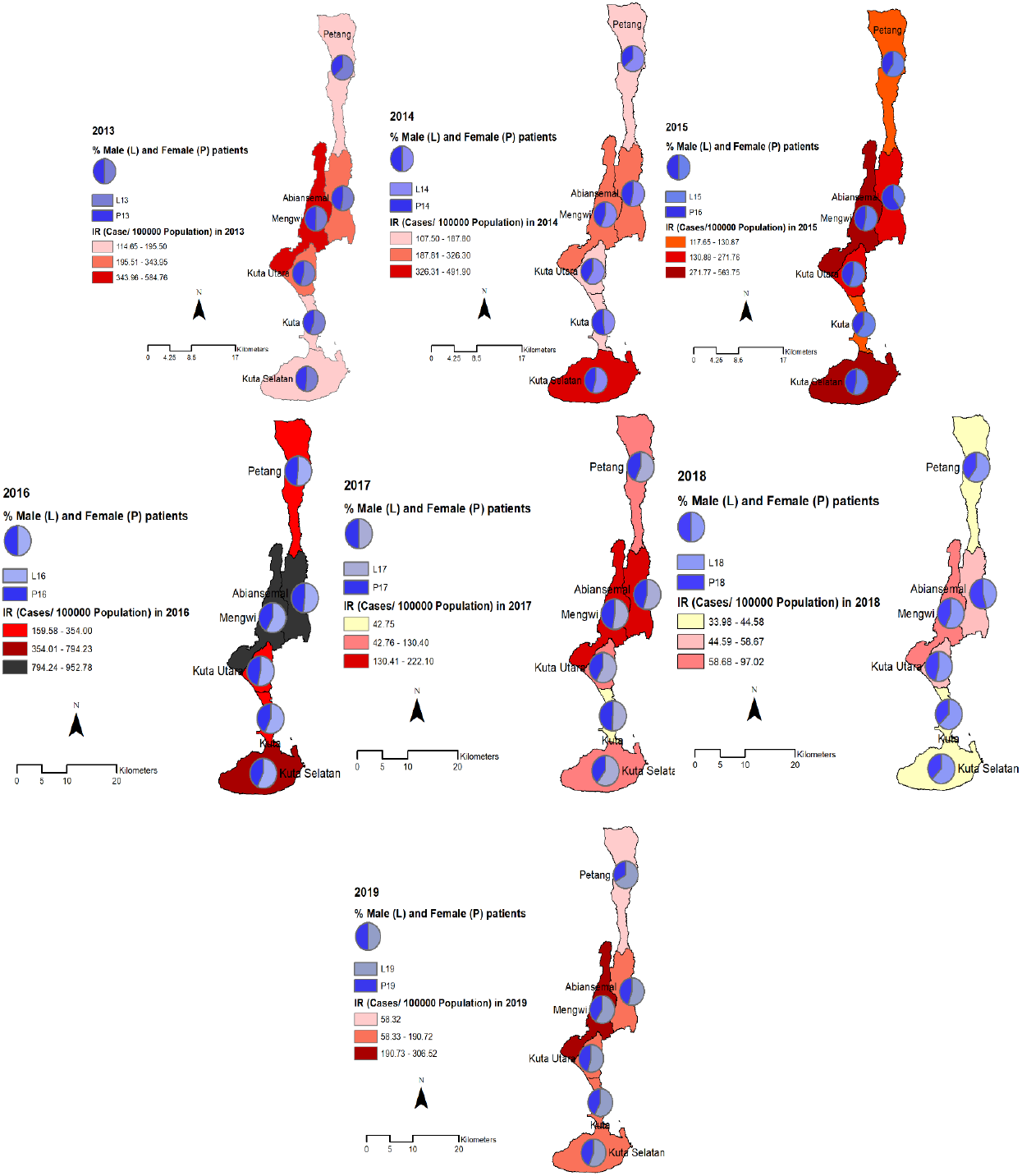
Geographical Distribution of Dengue Incidence Rates (IR) in 6 Sub-Districts in Badung 2013 – 2019.

The gender proportion of cases showed that during 7 years in 6 sub-districts were dominated by male patients, except in 2015 and 2018 for Abiensemal sub-disrict when the female patients were higher than male. However the rest of 5 illustrated the male patients were higher or slightly equal to female patient [Figure 1]. The detail table distribution of DF IR and gender proportion available in appendix 2.

### 3.2 Descriptive Statistics

During 60 months of observation (January 2013 – December 2017) there were 10,689 new DF cases. The maximum number of cases occurred in April 2016 and the minimum occurred in August 2017, with average of incidence rates almost 30 cases per 100000 Population. The highest temperature was recorded in November 2015 and the lowest was in August 2015 with average 27°C. The precipitation reached its peak in December 2017 (above 620 mm), while the lowest was in October 2015 (0 mm) with average 154.1 mm. The highest humidity was recorded in February 2016 and the lowest was in September 2014 with average value 79.98 percent. The highest number of local visitors was above 94 thousands recorded in November 2016 with average was nearly 50 thousands each month. While the highest foreign visitors was close to 600 thousands as recorded in August 2017 with average above 359 thousand every month. The detail statistic description is available in Table 1.

**Table 1.** Descriptive Frequency Distribution of Monthly Variables and Covariates 2013-2017.

| Variable and Covariates | Median | Mean | Min | Max | Standar Deviation |
| --- | --- | --- | --- | --- | --- |
| Case | 140,5 | 178,15 | 27 | 796 | 149,25 |
| IR | 23.1 | 29,26 | 4,2 | 128,2 | 24,23 |
| Temperature | 27.5 | 27,41 | 25,6 | 29 | 0,81 |
| Precipitation | 102.5 | 154,09 | 0 | 620,1 | 157,1962 |
| Humidity | 80 | 79,98 | 75 | 85 | 2,213 |
| Foreign Visitors | 344,510.5 | 359047,80 | 232935 | 593.337 | 84372,31 |
| Local Visitors | 47,682.5 | 48893,60 | 15.224 | 94.847 | 16761,33 |

The decomposition plot of DF time series data demonstrated an increasing trend that reached its highest on mid year of 2016 before it was sloping down thereafter. The plot also illustrated profound seasonal shape with consistent peaks around early months and the lowest around October - November for every year [Figure.2].

**Figure 2:**
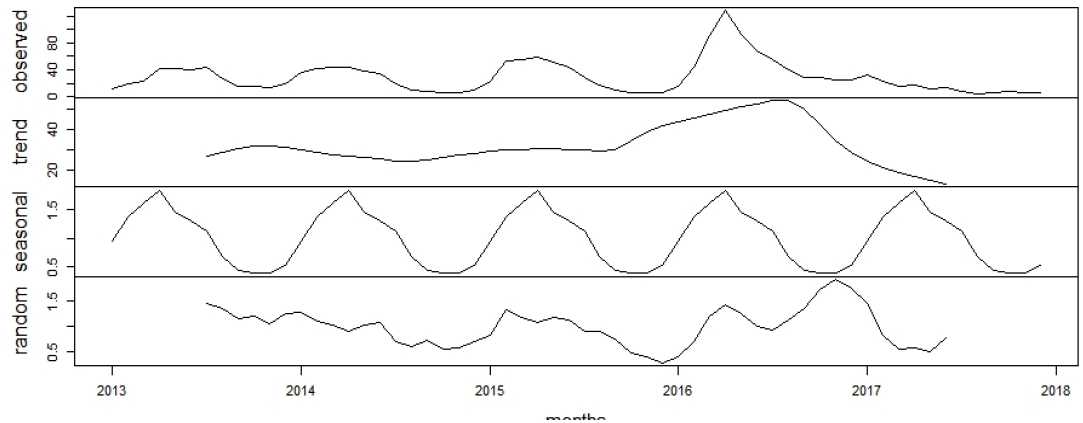
Decomposition of DF (IR/ 100000 Population) time series data in Badung District.

The box plots demonstrate the mean values distribution of variables in each 5 consecutive years. The highest mean of DF cases and incidence rates (IR) were reported in 2016. This pattern is similar to temperature variable, as the mean temperature in 2016 was reported as the highest. The rest of climatic variable have different pattern, whereas the mean of humidity in 2017 indicated as the highest, while the mean of precipitation in 2013 showed as the highest. The highest mean value of visitors was appeared in 2016 (local visitors) and 2017 (foreign visitors) [Figure. 3].

**Figure 3:**
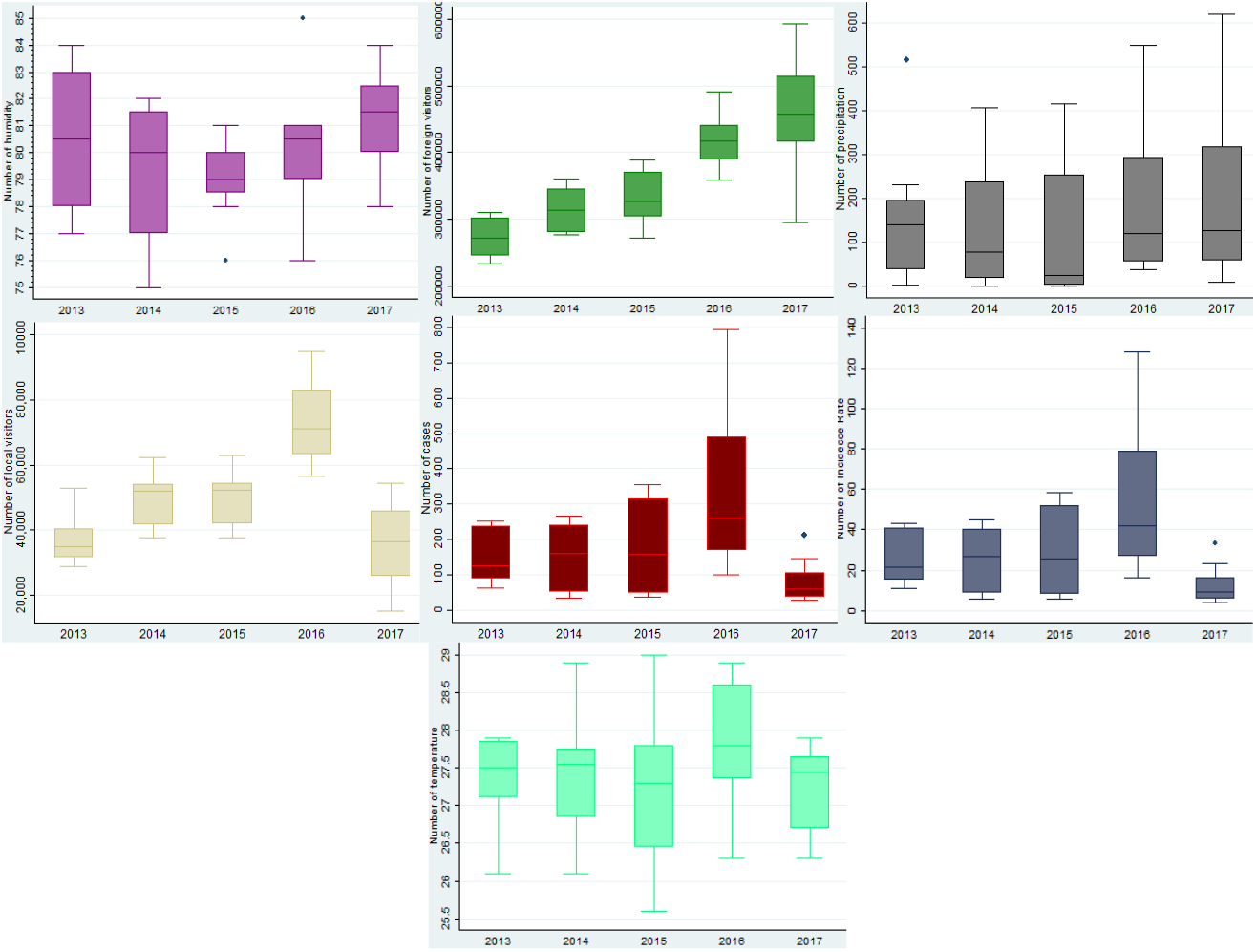
The monthly average of case, IR, temperature, precipitation, humidity, local and foreign visitors.

### 3.3 Analysis and Model Construction with Poisson Regression

The association between outcome and each of explanatory variables for multiple lags (0- 3 months lag) have been examined and all of them have significant result except for humidity at lag 3 months. Based on the AIC, *p value* and 95% CI there were 5 variable selected: temperature lag 3 months, precipitation lag 2 months, humidity lag 2 months, local visitors lag 1 month and foreign visitors lag 3 months. The detection of multicollinearity using Variance Inflation Factor (VIF) showed there is no value exceeding 4 means there is no multicollinearity among selected variables [Appendix 1].

DF cases is significantly associated with temperatures, the number of foreign tourists in the previous 3 months and local toruist in the previous 1 month. The DF Cases is also significantly associated with the precipitation and humidity occurring in the previous 2 months. DF cases increases by 5.909 × 10^−1^ (95% CI: 5.610 × 10^−1^ : 6.209 × 10^−1^), by 5.779 × 10^−4^ (95%CI: 3.882 × 10^−4^ : 7.676 × 10^−4^), 1.434 × 10^−2^ (95% CI: 0.303 × 10^−2^ : 2.565 ×10^−2^) and by 7.40 × 10^−6^ (95%CI: 5.88 × 10^−6^ : 8.91 × 10^−6^) for each increment 1°C of temperature (3 months lag), 1 mm of precipitation (2 months lag), 1% humidity (2 months lag) and additional 1 local visitor (1 month lag) respectively. However, the DF cases decreased by 2.18 × 10^−6^ (95% CI: - 2.50 × 10^−6^: −1.87 × 10^−6^) for every additional 1 foreign visitors in 3 months lag [Table 2].

**Table 2.** Result of Multivariate preliminary analysis of selected variables using poisson regression.

| Model with Selected Variables | Coefficient | P value | Confidence Interval(95% CI) |
| --- | --- | --- | --- |
| $\alpha$ (Intercept) | -11.98101 | 0,000 | -13.19878 : -10.76324 |
| Average Temperature lag3 | 0.59095 | 0,000 | 0.5610301 : 0.6208698 |
| Average Precipitation lag2 | 0.0005779 | 0,000 | 0.0003882 : 0.0007676 |
| Average Humidity lag2 | 0.0143432 | 0,013 | 0.0030307 : 0.0256557 |
| Local Visistors lag1 | $7.40 \times 10^{-6}$ | 0,000 | $5.88 \times 10^{-6} : 8.91 \times 10^{-6}$ |
| Foreign Visitors lag3 | $-2.18 \times 10^{-6}$ | 0,000 | $-2.50 \times 10^{-6} : -1.87 \times 10^{-6}$ |

## 4. Discussion

Climatic factors are found to have significant association with dengue cases in Badung District. Many possible mechanisms between climate and dengue were proposed by previous studies. Rising temperatures due to climate change led to shorter incubation period of mosquitoes, it impacted on the rapid and faster reproduction of the dengue vectors. Furthermore, optimal air temperature can accelerate mosquitos’ growth and the incubation period of the DENV virus (Huang et al. 2013, Tjaden et al. 2013, Zhang et al. 2016). Temperature ranges 25-27°C is optimal for the multiplication of mosquito vectors that it could change the pattern of interaction between infection diseases agent and human as the host (Goindin et al. 2015) as examined by previous studies about dengue fever transmission and environmental climate factors in Indonesia (Hasanah and Susanna 2018).

The optimum air humidity ranges from 70-80 percent which makes mosquitoes mobility more aggressive and increases the risk of viral transmission and the incidence of DHF disease. This association was signified by several studies (Astuti et al. 2019, Hasanah and Susanna 2018). This finding is also in line with the current study result in Badung district. However there was one study in South Sulawesi, Indonesia found otherwise stating that there is no significant influence of humidty on the lowering or increased incidence of DHF (Tosepu et al. 2018). The discrepancy is probably due to humidity variation in different topography of observed areas, moreover the location of South Sulawesi is closer to equator than Badung, Bali that it could drives different humidity characteristics and impacts.

Precipitation is one of the affecting factors for DF case in Badung district. The exploratory analysis for 0 – 3 months lags showed significant positive association. Similar result was reported from Neighbour country Thailand that precipitation together with temperature were indicated as the primary factors to drive dengue cases incidence (Langkulsen et al. 2020) as the presence of suitable precipitation could affects the humidity and initiates of mosquito-breeding place, as *Aedes aegypti* mosquitoes have high durability against high precipitation intensity. This finding also agrees with the study in Kerala, India that dengue occurrence during 2010 – 2014 was significantly associated with various lags of climatic factors (temperature, rainfall and humidity) and it showed spatial and temporal clustered (Valson and Soman 2017). However, the different result was reported by study in Singapore that found significant negative association between weekly percipitation and dengue incidence (Koh et al. 2018). The difference might be caused by the different time unit analysis as Badung study used monthly data while Singapore study used weekly data.

The number of local and foreign tourists is the influencing factor of DF case in Badung district. The result implies the association between the local and foreign visitors to the incidence of DF. This findings agrees with the previous study stated that dengue transmission was accounted for 2 percent of all of diseases experienced by visitors returned from endemic areas such us Southeast Asia (Wilder-Smith 2012). This result is also supported by the research in South Korea to found that in majority of imported DF cases were correlated with overseas travellers (Cho et al. 2018).

Aside from the geographical originis of Indonesia as tropical area located in the equator that it is conducive to support the breeding of *Aedes aegypti* as a vector of the dengue virus. Dengue transmission in Indonesia could be exacerbating by increasing population mobility and uncontrolled urbanization flows. Study in 2010 concluded that mobility of local and foreign visitors to Bali was significantly contributed for the DF transmission, in particularly densely populated of Badung district (Ishikawa and Shimogawara 2019). It is supported by another study in Japan that the dengue case (genotype I DENV-1) found in Okinawa in 2014 was confirmed as imported from Bali, Indonesia (Saito et al. 2016). These findings magnify the urgency of strengthening the monitoring and protection of international border starting from the arrival. Study in Yogyakarta reported that the breeding sites for *Aedes spp* is higher in airport leads to the potential for DF outbreak (Satoto et al. 2018). Nevertheless, the local residents and foresign vistors have been proven to have significant contribution to involve in dengue fever reduction program (Yoshikawa and Kusriastuti 2013).

The highly seasonal pattern of DF incidence found in Badung district agrees with the previous study of entire Bali Island (Dhewantara et al. 2019) also similar to the pattern of DF cases found in Timor Leste located in Timor Island (Wangdi et al. 2018). As the neighbors’s Island of Bali, this phenomena is most likely caused by the similar climate condition of southern part of equator that leads to the similar DF incidence pattern. The seasonal pattern was coincided with the climatic dynamics, for instance the peaks always occurred around early months, and this condition is concurrent with the high intention of reported precipitation in Badung that occurred around January – March. While, the lowest curve occurred around August – November that is coincided with the lowest reported humidity in this district.

The maps show that Abiansemal and Mengwi sub district frequently have highest dengue incidence from 2013 – 2019. According to Badung health profile, that these two sub-districts have the highest number of household: 39,663 and 22,124 respectively (PHO 2019). It could drives domestic transmission setting of DF incidences. While among 6 sub districts, Petang was consistenly has the lowest IR during 7 years. It could be due to its elevation is the highest in Badung district around 2,075 meters above sea level, as reported by Badung Bureau of Statistics (BPS 2016) that lead to the unafovourable weather, temperature and humidity for mosquitoes vectors’ proliferation. The DF cases were dominated by male patients in almost all districts during 2013 – 2019, most probably because in Badung district in majority male works as informal workers close to their house where water containers for mosquitoes breeding sites are available.

The present study have numbers of areas to be improved such as the inconsistent range of data and spatial units of analysis. For statistical analysis only covered 60 months (January 2013 – December 2017) at only at district level due to limited access to surveillance data. While for the visual geographical distribution, even though it covered 7 years (2013 – 2019) period however the spatial unit was only limited to 6 sub-districts that made it impossible to explore any further spatial pattern and analysis such as cluster and association. This study also has not integrated others potential risk variables into analysis such as health system and service accessibility, vector and larvae density, local population behaviour, population movement, transportation, interventions and others factors that directly and indirectly impacts on DF distribution.

Aside from the limitations stated, this study try to provide analysis that integrating both environmental (climate) and social determinant (visitors) in determining the risk for dengue spread. The result of this study could be the baseline information for developing comprehensive analysis on the impact of human movement on the emerging of infectious diseases.

## 5. Conclusion

The climate and tourism factors are the influencing factors for DF transmission dynamics in Badung District, therefore it need to be integrated for determining for DF monitoring and mitigation intervention in densely populated destination area.

## Data Availability

All data produced are available online at https://doi.org/10.7910/DVN/OLFCL1

## Acknowledgement

The authors thanks to the Diseases Prevention and Control Division of Bali Province Health Office, Bureau of Meteorology, Climatology, and Geophysics (BMKG) and Badung Regency Statistics Bureau (BPS) that have supported the research and article by providing the data.

## APPENDIX

**Appendix 1:** Result of Univariate preliminary analysis of selected variables using poisson regression and VIF identification using multivariate regression.

| Variables | VIF | IRR | 95% CI | P value<br>(poisson) | AIC | BIC |
| --- | --- | --- | --- | --- | --- | --- |
| **Without lag** |  |  |  |  |  |  |
| Average Temperature | 1,14 | 1.332228 | 1.300285 : 1.364955 | 0,000 | 6198.693 | 6202.881 |
| Average Precipitation | 1,88 | 0.9995045 | 0.9993775 : 0.9996316 | 0,000 | 6684.254 | 6688.443 |
| Average Humidity | 1,80 | 1.044416 | 1.035398 : 1.053513 | 0,000 | 6647.66 | 6651.849 |
| Local Visistors | 1,18 | 1.00001 | 1.000008 : 1.000011 | 0,000 | 6452.497 | 6456.686 |
| Foreign Visitors | 1,18 | 0.9999994 | 0.9999992 : 0.9999996 | 0,000 | 6717.178 | 6721.366 |
| **With lag1** |  |  |  |  |  |  |
| Average Temperature lag1 | 1,16 | 1.581323 | 1.542961 : 1.620639 | 0,000 | 5257.154 | 5261.309 |
| Average Precipitation lag1 | 1,83 | 1.001175 | 1.001053 : 1.001296 | 0,000 | 6293.349 | 6297.504 |
| Average Humidity lag1 | 1,75 | 1.098213 | 1.088593 : 1.107919 | 0,000 | 6197.537 | 6201.692 |
| Local Visistors lag1* | 1,18 | 1.000011 | 1.000010 : 1.000012 | 0,000 | 6257.705 | 6261.86 |
| Foreign Visitors lag1 | 1,17 | 0.9999984 | 0.9999981 : 0.9999986 | 0,000 | 6445.778 | 6449.933 |
| **With lag2** |  |  |  |  |  |  |
| Average Temperature lag2 | 1,15 | 1.774695 | 1.731259 : 1.819220 | 0,000 | 4456.405 | 4460.526 |
| Average Precipitation lag2* | 1,77 | 1.002338 | 1.002221 : 1.002455 | 0,000 | 5190.697 | 5194.818 |
| Average Humidity lag2* | 1,70 | 1.100248 | 1.090501 : 1.110083 | 0,000 | 6154.343 | 6158.463 |
| Local Visistors lag2 | 1,18 | 1.00001 | 1.000009 : 1.000011 | 0,000 | 6271.487 | 6275.608 |
| Foreign Visitors lag2 | 1,17 | 0.9999978 | 0.9999975 : 0.999998 | 0,000 | 6263.738 | 6267.858 |
| **With lag3** |  |  |  |  |  |  |
| Average Temperature lag3* | 1,17 | 2.084046 | 2.032162 : 2.137255 | 0,000 | 3128.901 | 3132.987 |
| Average Precipitation lag3 | 1,77 | 1.002119 | 1.002001 : 1.002238 | 0,000 | 5442.098 | 5446.184 |
| Average Humidity lag3 | 1,70 | 0.9962442 | 0.9874804 : 1.005086 | 0.404 | 6577.703 | 6581.789 |
| Local Visistors lag3 | 1,19 | 1.00001 | 1.000009 : 1.000011 | 0,000 | 6261.01 | 6265.096 |
| Foreign Visitors lag3* | 1,19 | 0.9999973 | 0.9999971 : 0.9999976 | 0,000 | 6105.728 | 6109.814 |
| **With selected lag** |  |  |  |  |  |  |
| Average Temperature lag3 | 1,51 | 2.084046 | 2.032162 : 2.137255 | 0,000 | 3128.901 | 3132.987 |
| Average Precipitation lag2 | 2,07 | 1.002338 | 1.002221 : 1.002455 | 0,000 | 5190.697 | 5194.818 |
| Average Humidity lag2 | 1,74 | 1.100248 | 1.090501 : 1.110083 | 0,000 | 6154.343 | 6158.463 |
| Local Visistors lag1 | 1,38 | 1.000011 | 1.000010 : 1.000012 | 0,000 | 6257.705 | 6261.86 |
| Foreign Visitors lag3 | 1,20 | 0.9999973 | 0.9999971 : 0.9999976 | 0,000 | 6105.728 | 6109.814 |

**Appendix 2:** Distribution of DF in 6 Sub Districts in Badung from 2013 - 2019.

| Sub-Districts | 2013 |  |  | 2014 |  |  | 2015 |  |  | 2016 |  |  | 2017 |  |  | 2018 |  |  | 2019 |  |  |
| --- | --- | --- | --- | --- | --- | --- | --- | --- | --- | --- | --- | --- | --- | --- | --- | --- | --- | --- | --- | --- | --- |
|  | Male (%) | Female (%) | IR/100000 Population | Male (%) | Female (%) | IR/100000 Population | Male (%) | Female (%) | IR/100000 Population | Male (%) | Female (%) | IR/100000 Population | Male (%) | Female (%) | IR/100000 Population | Male (%) | Female (%) | IR/100000 Population | Male (%) | Female (%) | IR/100000 Population |
| Abiansemal | 52.90 | 47.10 | 343.95 | 53.96 | 46.04 | 292.70 | 40.08 | 59.92 | 271.76 | 51.96 | 48.04 | 923.53 | 54.68 | 45.32 | 221.50 | 46.30 | 53.70 | 58.67 | 54.55 | 45.45 | 142.95 |
| Kuta | 55.96 | 44.04 | 114.65 | 47.62 | 52.38 | 107.50 | 60.17 | 39.83 | 117.65 | 57.32 | 42.68 | 159.58 | 62.22 | 60.00 | 42.75 | 62.50 | 37.50 | 44.58 | 57.50 | 42.50 | 145.32 |
| Kuta Selatan | 52.09 | 47.91 | 195.50 | 54.41 | 45.59 | 491.90 | 54.36 | 45.64 | 563.75 | 57.26 | 42.74 | 794.23 | 60.20 | 39.80 | 126.76 | 62.50 | 37.50 | 33.98 | 55.89 | 44.11 | 173.85 |
| Kuta Utara | 54.68 | 45.32 | 294.70 | 58.67 | 41.33 | 187.80 | 55.45 | 44.55 | 259.65 | 53.44 | 46.56 | 354.00 | 57.89 | 42.11 | 130.40 | 54.29 | 45.71 | 51.96 | 54.92 | 45.08 | 190.72 |
| Mengwi | 50.20 | 49.80 | 584.76 | 56.22 | 43.54 | 326.30 | 53.80 | 46.20 | 489.62 | 57.79 | 42.21 | 952.78 | 52.58 | 47.42 | 222.10 | 56.25 | 43.75 | 97.02 | 58.97 | 41.03 | 306.52 |
| Petang | 62.86 | 37.14 | 133.79 | 63.89 | 36.11 | 138.00 | 58.82 | 41.18 | 130.87 | 51.69 | 48.31 | 343.50 | 56.67 | 43.33 | 116.10 | 60.00 | 40.00 | 38.82 | 66.67 | 33.33 | 58.32 |

